# Challenging the gold standard: critical limitations in clinical detection of drug-resistant tuberculosis

**DOI:** 10.1101/2023.02.27.23286518

**Authors:** Sarah N. Danchuk, Ori E. Solomon, Thomas A. Kohl, Stefan Niemann, Dick van Soolingen, Jakko van Ingen, Joy S. Michael, Marcel A. Behr

**Affiliations:** Department of Microbiology and Immunology, McGill University; Infectious Diseases and Immunity in Global Health Program, Research Institute of the McGill University Health Centre; McGill International TB Centre; Molecular and Experimental Mycobacteriology, Research Center Borstel; German Center for Infection Research, Hamburg-Lübeck-Borstel-Riems, Germany; National Institute for Public Health and Environment, The Netherlands; Department of Medical Microbiology, Radboudumc Center for Infectious Diseases, Radboud University Medical Center, Nijmegen, the Netherlands; Christian Medical College, Vellore, India; Department of Medicine, McGill University Health Centre

**Keywords:** Quality control, mycobacterial laboratory, antibiotic resistance, BCG, Whole Genome Sequencing, heteroresistance

## Abstract

Heteroresistant infections - defined as infections in which minority drug-resistant (DR) populations are present - are a challenge in infectious disease control. In *Mycobacterium tuberculosis*, heteroresistance poses challenges in diagnosis and has been linked with poor treatment outcomes. We compared the analytic sensitivity of molecular methods, such as GeneXpert and whole genome sequencing (WGS) in detecting heteroresistance when compared to the ‘gold standard’ phenotypic assay: the agar proportion method (APM). Using defined mono-resisitant BCG strains we determined the limit of detection (LOD) of rifampin-R (RIF-R) detection was 1% using APM, 60% using Xpert MTB/RIF and 10% using Xpert MTB/RIF Ultra. To evaluate clinical WGS pipelines, a blinded panel of BCG mixtures was sent to 3 clinical labs. These were composed of either a) RIF-R plus isoniazid-R (INH-R) BCG or b) fluoroquinolone-R (FQ-R) plus clofazimine-R/bedaquiline-R (CLZ/BDQ-R) BCG. No labs called resistance at 1%; all labs called RIF-R at 10% or greater and two out of three labs reported FQ-R at 10%. Two labs were able to detect the majority population (either INH-R or CLZ/BDQ-R) at 50%. Importantly, where labs did not report resistance in the majority population, the mutations were present in the raw data but excluded from the final analysis. In conclusion, the gold standard APM more reliably detects minority resistant populations than molecular tests. Further research is required to determine whether the higher LOD of molecular tests is associated with deleterious patient outcomes and the potential effects on transmission of resistance at the population level.

## Introduction

Tuberculosis (TB) is the second leading cause of mortality by a single infectious agent and the thirteenth leading cause of death globally^1^. Eradication efforts have been largely hindered by the emergence of drug-resistant *Mycobacterium tuberculosis* (*M. tb*), the causative agent of TB^1^. In 2021, there were an estimated 450,000 cases of rifampicin-resistant TB/multidrug resistant TB (RR-TB/MDR-TB) and 119,000 deaths attributed specifically to this^1^. The cornerstone of DR-TB management is detection as inadequate treatment can result in failure to cure at the individual level and onward propagation of DR-*M. tb* (DR-MTB) isolates to their contacts^2^.

Since the 1960s, the reference method for detecting DR-MTB has been phenotypic testing by the agar proportion method (APM)^3^. The APM was developed on the premise that when a certain proportion of bacilli is resistant to the antibiotic, clinical success is unlikely. This quantitative property has been operationally reduced into a dichotomous result (resistant or susceptible) based on a threshold of 1% resistance^4^. Patients in whom more than 1% of the *M. tb* population grow in the presence of the antibiotic are not expected to respond to treatment with that antibiotic as the resistant fraction is expected to dominate within several doubling times following initiation of treatment. The use of a 1% threshold in the *M*.*tb* world contrasts with a 0.1% threshold used in other sections of the microbiology lab; presumably resistant bacteria at less than 1% can be managed by multi-drug therapy^4,5^.

Culture-based DSTs are difficult and costly to implement and present a challenge of turnaround time, where clinicians aim to start therapy promptly for new diagnosed patients. To mitigate these challenges, molecular tests have been implemented globally for 1) the detection of *M. tb* and 2) the determination of first- and second-line drug susceptibility^6^.These tests span from simple tests used in peripheral labs (e.g., the GeneXpert MTB/RIF assay; Cepheid, Sunnyvale CA, USA) to Whole Genome Sequencing-based prediction of resistance^7^. Given the importance of molecular testing for DR-MTB detection, our lab generated a panel of first- and second-line mono-resistant *M. bovis* BCG strains to serve as quality controls for both phenotypic and genotypic testing^8^. The use of these strains to validate the detection of *M. tb* in a peripheral setting has previously been described^8^. In this study we sought to evaluate the capacity of molecular assays to detect heteroresistance, using defined mixtures of these mono-resistant BCG strains.

## Methods

### Heteroresistant samples

The development of mono-resistant BCG strains has been previously published^8^. As the agar proportion method (APM) detects resistant organisms at 1% of the bacterial population^4,9^, we created mixtures of BCG strains at different ratios, to test detection at 50%, 10% or 1%. To validate our findings against published observations about the Xpert MTB/RIF^10,11^, we also generated a 60% RIF-R mixture (Table 1).

**Table 1.**
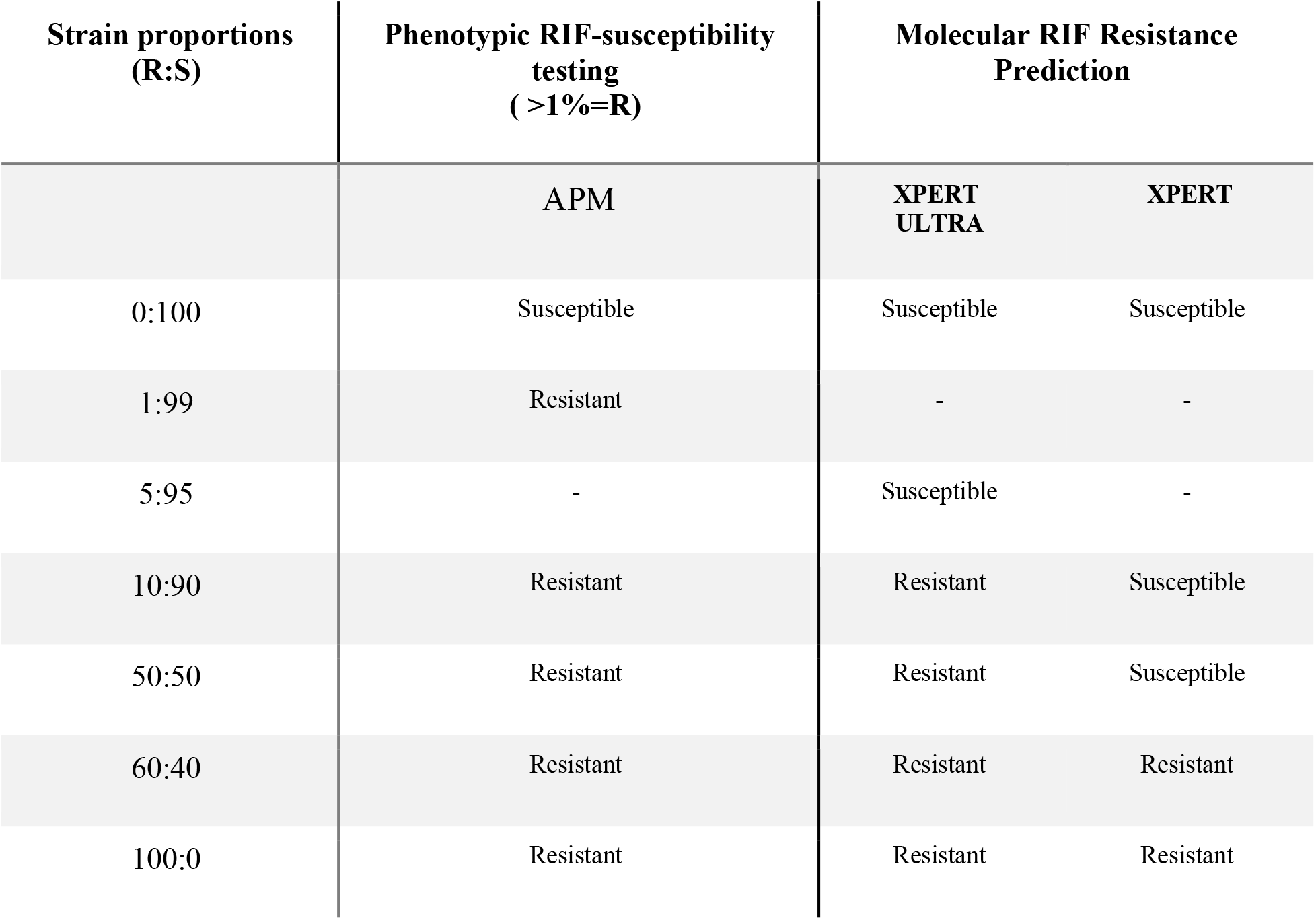
Phenotypic and molecular detection of RIF-heteroresistance. ‘Resistant’ defined as ≥ 1% growth in antibiotic media compared to antibiotic free media. -: DST platform not tested at this proportion.

### Agar proportion method (APM)

Wildtype (WT) *M. bovis* BCG and RIF-R *M. bovis* BCG (RpoB S531L) were grown in 7H9 complete (0.5% glycerol, 0.1% Tween, 10% Albumin Dextrose Catalase (ADC) supplement) to log phase (OD_600_ 0.5-1). WT cultures were grown in the absence of antibiotics whereas RIF-R BCG was grown in the presence of 1 ug/mL RIF. Cultures were adjusted to 0.5 MacFarland standard using 7H9 Middlebrook complete media and serially diluted (10^−2^ and 10^−4^). 0.1 mL of culture mixes were inoculated on 7H10 Oleic Acid Albumin Dextrose Catalase (OADC) quadrant petri dishes containing: 1) no antibiotics, 2) 1 ug/mL Levofloxacin, 3) 1 ug/mL INH, and 4) 1 ug/mL RIF quadrants^4^. Cultures were incubated at 37°C for three weeks and CFUs were counted. Resistance was defined as CFUs on antibiotic quadrant ≥1% of antibiotic-free quadrant^4^.

### Xpert MTB/RIF + Xpert MTB/RIF Ultra

For both Xpert MTB/RIF and Xpert MTB/RIF Ultra (hereby referred to as Xpert and Xpert Ultra, respectively), assays were performed per manufacturer instructions (Cepheid, Sunnyvale, CA USA). Briefly, 0.5 mL of sample + 1.5 mL of Sample Reagent (SR, included with assay) was aliquoted into 15 mL tube, vortexed for 10 seconds and incubated at room temperature (RT) for 10 minutes. The sample was then vortexed for an additional 10 seconds, incubated for 5 minutes at RT and added to the GeneXpert cartridge.

### gDNA extraction

gDNA was extracted from RIF-R (RpoB S531L), INH-R (KatG AA428del), FQ-R (GyrA D94G), and CLZ/BDQ-R (Rv0678c S63R) mono-resistant *M. bovis* BCG strains using the Van Sooligen protocol (previously described)^12^. Concentration (ng/uL) was measured using Quant-iT™ PicoGreen™ dsDNA assay per manufacturer protocol (Life Technologies Corporation, Eugene, Oregon USA). gDNA was prepared in the following proportions of 50:50, 10:90, and 1:99 (Table 2) and sent to the testing labs, blinded to sample identity, for both first- and second-line antibiotic assessment. Heteroresistant mixtures were then evaluated by WGS using laboratory-specific bioinformatic pipelines for isolate characterization and drug susceptibility. Reports were returned and interpreted internally.

**Table 2.** Resistance calling following WGS of blinded samples. Samples evaluated were composed of 50:50, 10:90 and 1:99 RIF-R:INH-R gDNA to determine LOD of clinical sequencing. 100% RIF-R gDNA and 100% SM-R gDNA were included as a control for all labs.

| Labs | RIF-R:INH-R proportions |  |  |  |  |  |
| --- | --- | --- | --- | --- | --- | --- |
|  | RIF-R | INH-R | RIF-R | INH-R | RIF-R | INH-R |
|  | 50% | 50% | 10% | 90% | 1% | 99% |
| **<u>A</u>** | Detected | Detected | Detected | Detected | Not detected | Detected |
| **<u>B</u>** | Detected | Detected | Detected | Detected | Not detected | Detected |
| **<u>C</u>** | Detected | Not Detected | Detected | Not Detected | Not detected | Not detected |

## Results

Minority populations at the limit of 1% resistance were detected using the phenotypic assay (APM) (Figure 1). As has been previously published, Xpert was unable to accurately detect RIF-R at 50%^10,11^. To verify the threshold of detection, the percent of RIF-R bacteria present within the sample was increased, confirming that the assay can detect RIF-R at a proportion of ≥60%. When the same samples were run using Xpert Ultra, minority populations were detected at proportions ≥10%. When this was further diluted to 5%, the Xpert Ultra failed to detect the minority population (Table 1).

**Figure 1.**
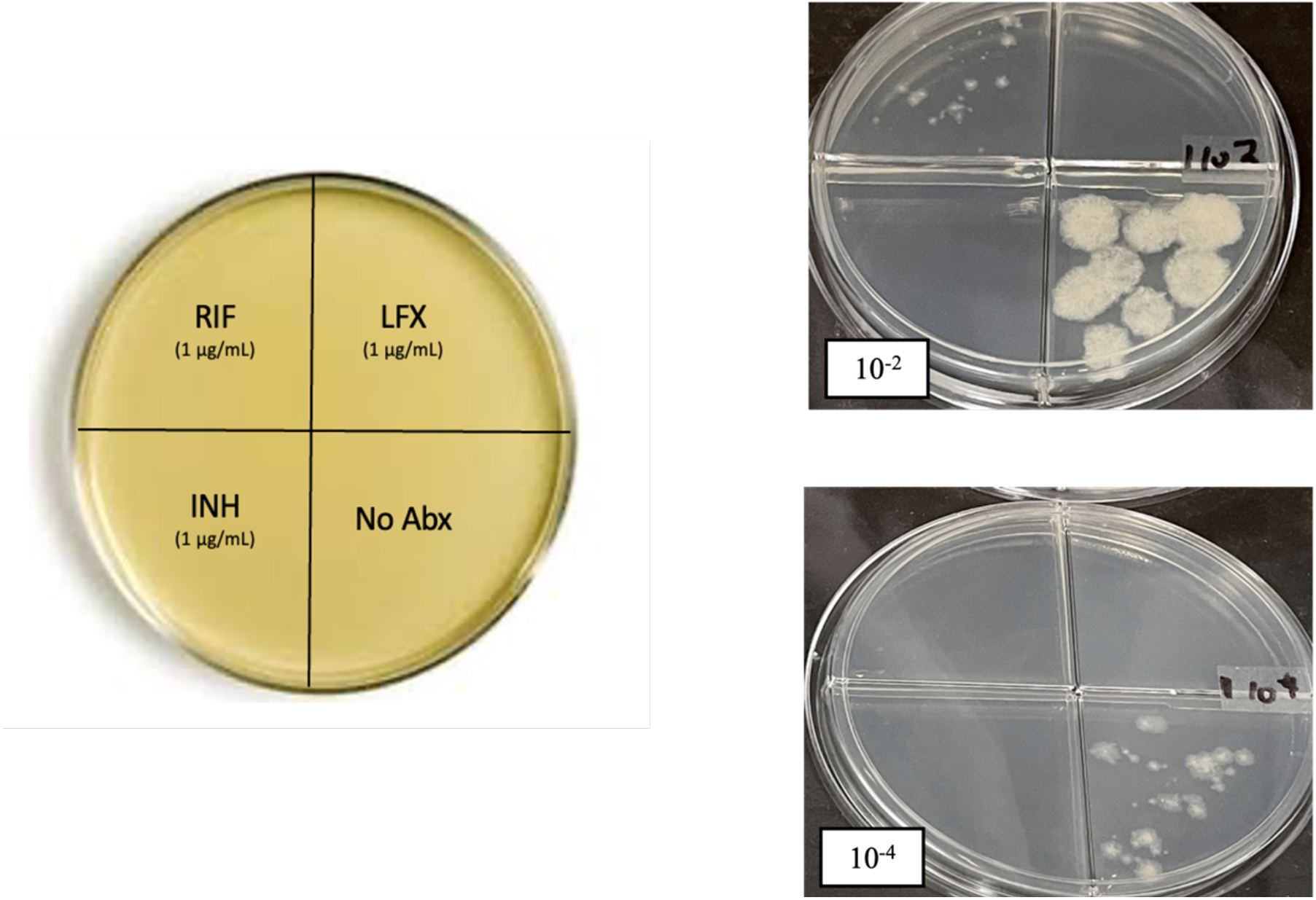
**Left:** schematic of quadrant plate used in lab for phenotypic detection. INH (1 μg/mL) and LFX (1 μg/mL) were used as no-growth controls (100% drug susceptibility) whereas the no antibiotics was used as a viability control (100% growth). **Right:** BCG RIF-R (RpoB S531L) and WT strains were prepared in a 1:99 mixture. Cultures were resuspended to 0.5 McFarland standard, serial diluted to 10-2 and 10-4, and plated on 7H10 agar + OADC, as per CLSI guidelines. LOD = 1% (n=4)

To evaluate the limit of detection (LOD) and capabilities of clinical WGS, we sent mixed mono-resistant populations to three reference labs. Samples were analyzed and a resistance report was returned as would be done to guide ‘patient’ treatment by local medical practitioners (Figure 2). All labs were consistently able to detect PZA resistance, characteristic of *M. bovis* BCG due to a mutation in *pncA*, in addition to 100% RIF-R and 100% streptomycin-R (SM-R) control strains. The RIF-R (RpoB S531L) strain was mixed with an INH-R strain (KatG del428 mutant) in the proportions described in Table 2. Two of the three labs were able to detect the INH-R mutant. The third lab detected the mutation in their variant calling but, this did not translate into the final resistance report. For RIF-R detection, all 3 labs detected minority RIF-R populations at 10% but none were able to detect at 1%. (Table 2).

**Figure 2.**
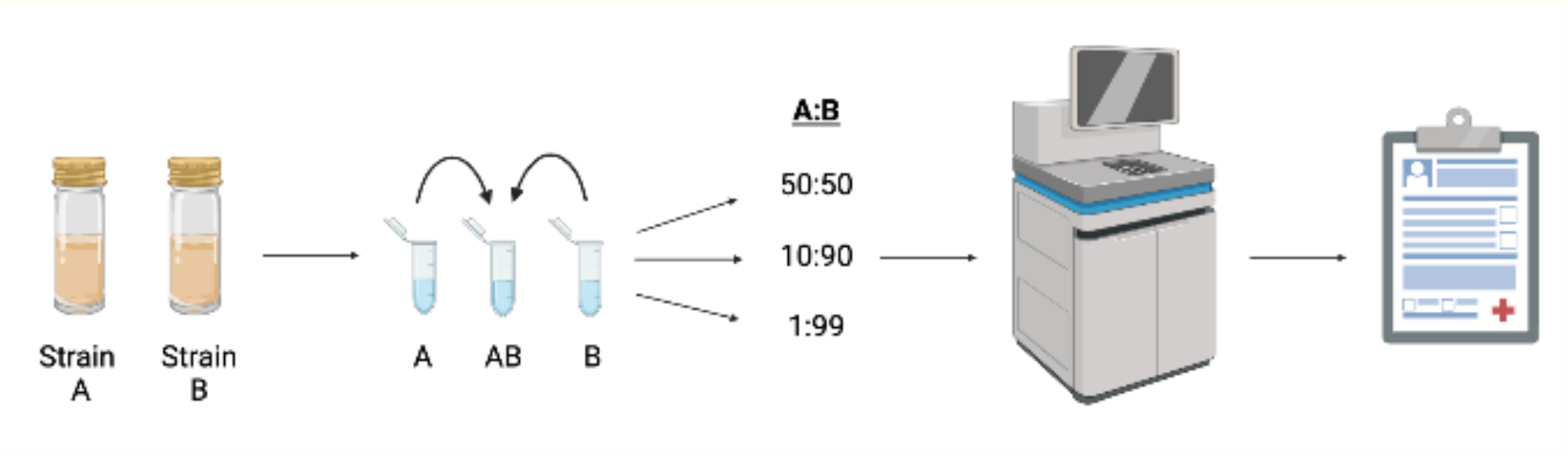
Preparation of heteroresistant strains and subsequent workflow. For first-line antibiotics ‘strain A’ =RIF-R BCG (S531L) and ‘strain B’=INH-R BCG (KatG428del). For second-line antibiotics ‘strain A’ =FQ-R BCG (GyrA D94G) and ‘strain B’ =CLZ/BDQ-R (Rv0678c S63R). For both first- and second-line antibiotics ‘strain A’ composed the minority population whereas ‘strain B’ was the majority. Following WGS and variant calling, resistance reports were generated, and drug susceptibility was determined. Figure created with BioRender.com

An attractive characteristic of clinical WGS is that numerous antibiotics can be assessed using a singular assay. We evaluated second-line resistance detection using the FQ-R (GyrA D94G) and CLZ/BDQ-R (Rv0678c S63R) strains described in Table 3. Concordant with results for first-line antibiotics, the LOD for second-line detection was 10%. Two out of three labs could accurately detect FQ-R at 10% of total sample, but not at 1%. One lab was unable to detect FQ-R at either 1% or 10%. Comparably, CLZ/BDQ-R was also accurately reported by two out of three labs. The third lab detected the mutation (Rv0678c S63R) in the variant calling but did not include it in the final resistance report at any proportion.

**Table 3.** Resistance calling following WGS of blinded samples. Samples evaluated were composed of 50:50, 10:90 and 1:99 FQ-R:CLZ/BDQ-R gDNA to determine LOD of clinical sequencing. 100% RIF-R gDNA and 100% SM-R gDNA were included as a control for all labs.

| Labs | FQ-R:CLZ/BDQ-R proportions |  |  |  |  |  |
| --- | --- | --- | --- | --- | --- | --- |
|  | FQ-R | CLZ/BDQ-R | FQ-R | CLZ/BDQ-R | FQ-R | CLZ/BDQ-R |
|  | 50% | 50% | 10% | 90% | 1% | 99% |
| <u>A</u> | Detected | Not detected | Detected | Not detected | Not detected | Not detected |
| B | Detected | Detected | Detected | Detected | - | - |
| <u>C</u> | Detected | Detected | Not detected | Detected | Not detected | Detected |

## Discussion

Adequacy of TB treatment hinges on effective and accurate diagnosis. Recently, heteroresistance and mixed infections have become a valid and rising concern in TB diagnosis and treatment^2^. Previous studies have shown that up to 20% tested clinical isolates contain heteroresistant populations, which is particularly concerning as these mixed populations may lead to treatment failure and subsequent DR-TB^13,14,15^. Without adequate diagnostics, patient care and public health may be compromised: not only will the patient be exposed to the adverse effects of ineffective antibiotics, but DR-MTB may be selected for due to inadequate treatment. Our results confirm that phenotypic testing, such as the APM, will be able to detect resistant populations down to 1%, whereas current molecular tests cannot. WGS offers the promise of detecting mutations across the complete genome, beyond the limited regions covered by probe-based assays such as the Xpert MTB/RIF assays. However, our results indicate that this method is also limited in its ability to detect resistant subpopulations.

Through this study we observed two main caveats to clinical WGS. First, if the in-house sequencing and analysis pipelines are not optimized to include allele frequencies below 10%, these variants will be filtered from analysis despite being present. New molecular tests with increased depth of coverage, such as the Deeplex assay, have been shown to lower the LOD to 5%^16^. However, variant calling at a lower frequency and read depth may lead to an increase of false positive calls due to contaminant species reads or sample cross-contamination^17,18^. Second, if the mutation is non-canonical (or otherwise absent from the laboratory resistance mutation catalogue), the sample may be reported as drug susceptible, as we observed for the KatG del428 strain. For the Rv0678c S63R mutant, the role of this mutation in BDQ is contested^19^ which highlights the need for studies using allelic exchange to ascertain which resistance-associated mutations are the cause of the phenotype. These findings emphasize the importance of constantly updating resistance mutation catalogues when using genotypic DST.

Clinicians supervising TB care and laboratories offering molecular testing should be aware of these analytic limitations. One potential solution is for reference laboratories to retain phenotypic testing capacity, for cases not responding to therapy and/or for a random sample of isolates, to ensure concordance of molecular predictions with phenotypic results. Regulators and developers of novel diagnostic tests must also consider the importance of detecting minority or emerging resistance populations, and the consequences of ignoring them. This is exemplified by the WHO’s “Target Product Profile for next-generation drug susceptibility testing for *M. tb* at peripheral centres”, in which the minimal requirement for minor variant detection is 20%^20^. The consequence of this adjustment is currently unknown. Though it is crucial to have patients started on antibiotic therapy as quickly as possible, it is also necessary for this therapy to be correct.

## Data Availability

All data are available for future analysis. All data produced in the present study are available upon reasonable request to the authors

## CONCLUSION

We demonstrated that phenotypic testing has a greater analytic sensitivity for detecting minority populations of resistant bacteria compared to molecular testing. The Xpert assay is still commonly used in many high-burden DR-MTB settings, despite having an LOD of 60%. Comparatively, we noted that the sensitivity of Xpert Ultra and WGS had similar LODs at around 10%. Further investigation of the correlation between phenotypic/genotypic testing, transmission of resistant strains, and treatment outcomes is needed. Finally, these blind spots in conjunction with the rise of drug resistance lead us to question the diagnostic cut-off being used in DST. With rates of DR-MTB rising worldwide, should the 1% threshold for resistance be revisited? Can we switch to tests that can only detect 10% resistant populations or should we be moving to a more stringent cut-off, as is used in other sectors of the microbiology lab, below 1%?

